# The impact of a gamified intervention on daily steps in real-life conditions: a retrospective analysis of 4800 individuals

**DOI:** 10.1101/2022.11.18.22282458

**Authors:** Alexandre Mazéas, Cyril Forestier, Guillaume Harel, Martine Duclos, Aïna Chalabaev

**Author notes:** **Data, code & supplemental materials:** https://osf.io/scnu7/.

## Abstract

**Background:** Digital interventions integrating gamification features hold promise to promote daily steps. However, results regarding the effectiveness of this type of intervention are heterogeneous and not yet confirmed in real-life contexts.

**Objective:** This study aimed to examine the effectiveness of a gamified intervention and its potential moderators in a large sample using real-world data. Specifically, we tested (1) whether a gamified intervention enhanced daily steps during the intervention and follow-up periods compared to baseline, (2) whether this enhancement was higher in participants to the intervention than in nonparticipants, and (3) what participants’ characteristics or intervention parameters moderated the effect of the program.

**Methods:** Data from 4819 individuals who registered for a mHealth Kiplin program between 2019 and 2022 were retrospectively analyzed. In this intervention, participants could take part in one or several games where their daily step count was tracked, allowing individuals to play with their overall activity. Nonparticipants are people who registered to the program but did not take part in the intervention and were considered as a control group. Daily step counts were measured via accelerometers embedded in either commercial wearables or smartphones of the participants. Exposure to the intervention, the intervention content, and participants’ characteristics were included in multilevel models to test the study objectives.

**Results:** Participants in the intervention group demonstrated a significantly greater increase in mean daily steps from baseline compared to nonparticipants (*p*<.0001). However, intervention effectiveness depended on participants’ initial physical activity. Whereas the daily steps of participants with <7500 baseline daily steps significantly improved from baseline both during the Kiplin intervention (+3291 daily steps) and during follow-up periods (+945 daily steps), participants with a higher baseline had no improvement or significant decreases in daily steps after the intervention. Age (*p*<.0001) and exposure (*p*<.0001) positively moderated the intervention effect.

**Conclusions:** In real-world settings and among a large sample, the Kiplin intervention was significantly effective to increase the daily steps of participants from baseline, during intervention and follow-up periods, compared to nonparticipants. Interestingly, responses to the intervention differed based on participant’ initial steps with the existence of a plateau effect. Drawing on the insights of the self-determination theory, we can assume that the effect of gamification could depend of the initial motivation and activity of participants.

## Introduction

Physically inactive individuals are at higher risk of developing non-communicable diseases – such as cardiovascular diseases, cancers, type 2 diabetes mellitus, or obesity – and mental health issues compared to the most active ones [1]. Yet, one-third of the world’s population is insufficiently active [2,3] and the trend is downward, with adults performing on average 1000 fewer steps per day than 2 decades ago [4]. Additionally, it has recently been reported that the global population step count did not return to pre-pandemic levels in the 2 years following the onset of the COVID-19 outbreak [5]. The number of steps per day is a simple and convenient measure of physical activity (PA). Recent research suggests that an increase in the daily step count is associated with a progressively lower risk of all-cause mortality. Walking an additional 1,000 steps per day can help reduce the risk of all-cause mortality [6]. For adults aged 60 years and older, this reduction in mortality rates is observed up to approximately 6000–8000 steps per day, while for adults under 60 years, the threshold is around 8000–10,000 steps per day [7]. However, sustaining this increase over time is crucial to achieve tangible health benefits [8]. Despite the efficacy of current programs in eliciting initial changes in individuals’ PA, they often struggle to induce long-term behavioral shifts in the behavior [9]. In this context, there is an urgent need to sustainably increase the number of daily steps of individuals in primary, secondary, and tertiary prevention.

Digital behavior change interventions are promising avenues to promote daily steps. Smartphones and digital tools, ubiquitous in our daily lives, offer several advantages, including their widespread availability, relatively low cost, and ability to access content quickly from anywhere [10–12]. Moreover, these technologies can collect real-time data in natural contexts (i.e., daily step counts can be measured via accelerometers embedded in either commercial wearables such as Fitbit or smartphones) and present it in quantified formats, providing opportunities for exploration and reflection. This facilitates the implementation of powerful behavior change techniques, such as goal setting and self-monitoring, potentially influencing behaviors [11]. However, there are concerns about the ability of digital programs to engage participants once the novelty wears off or to be effective on any type of audience regardless of their age, sociodemographic, or health status. In this context, gamification strategies introduce an exciting roadmap for addressing these challenges.

Gamification refers to the use of game elements in nongame contexts [13] and allows to transform a routine activity into a more engaging one. The self-determination theory (SDT) [14] is a commonly used theoretical framework for understanding the motivational impact of gamification on behavior. The SDT suggests the existence of different types of motivation that can be pictured on a continuum ranging from lack of motivation to completely autonomous motivation, in which the behavior comes from the individual’s will. On the opposite, controlled motivation will lead the individual to practice for the consequences that the activity can bring and not for the activity itself. The SDT holds that people will be more likely to perform the behavior in the long-term when their motivation is autonomous rather than controlled. Thus, autonomous forms of motivation represent more sustainable drivers of engagement and are an important predictor of the long-term maintenance of physical practice [15,16]. Autonomous motivation occurs when people perform an activity for their own satisfaction, inherent interest, and enjoyment. In addition, three basic psychological needs are presumed to achieve self-determination: the need for autonomy (i.e., need to feel responsible of one’s own actions), competence (i.e., need to feel effective in one’s interactions with the environment), and relatedness (i.e., need to feel connected to other people).

In addition to providing fun and playful experiences to users, gamification can effectively address basic psychological needs [17]. Firstly, gamification strategies such as point scoring, badges, levels, and competitions serve to sustain the need for competence by offering feedback on users’ behaviors. Secondly, customizable game environments or user choices can support autonomy. Finally, features such as leaderboards, team structures, groups, or communication functions can foster a sense of relatedness. From this perspective, a gamified intervention would feed the autonomous motivation of participants and would be more correlated with the long-term adherence to PA. However, from another perspective, several criticisms have been leveled at gamification including the fact that these mechanisms are reward-oriented and that, still in line with the SDT, the use of external rewards can reduce autonomous motivation [18,19].

A recent meta-analysis [20] revealed that digital gamified interventions, lasting on average 12 weeks, improved daily steps by 1600 steps on average. Importantly, the results showed that gamified interventions a) appear more effective than digital non-gamified interventions, b) seem appropriate for any type of user regardless of their age or health status, and c) the PA improvement persists after follow-up periods lasting in average 14 weeks, with a very small to small effect size. As a result, gamified interventions are emerging as interesting behavior change tools to tackle the physical inactivity pandemic. However, these findings obtained from randomized controlled trials do not always reflect what happens in real-life settings [21]. In addition, the effect sizes reported in this meta-analysis were heterogeneous and the authors found high between-study heterogeneity (e.g., *I*^*2*^ = 82%).

If this heterogeneity may be explained by differences in study quality or diversity of designs in the included studies, the behavior change intervention ontology proposed by Michie et al. [22] argues that heterogeneity in behavioral interventions could also be explained by different variables such as intervention characteristics (e.g., content, delivery), the context (e.g., characteristics of the population targeted such as demographics, setting such as the policy environment or physical location), exposure of participants with the program (e.g., engagement and reach), and the mechanisms of action (the processes by which interventions influence the target behavior). Considering these variables within gamification contexts could provide a useful means to better understand the conditions under which interventions are successful. Furthermore, based on the SDT, we can envisage that gamification techniques will not have the same impact on all users, depending on their initial motivation and the way they perceive games.

The present study investigated these questions based on a retrospective analysis of real-world data collected from a large sample of adult participants who were proposed a mHealth gamified intervention developed by the company Kiplin in France from 2019 to 2022. In this one, participants could take part in one or several collective games where their daily step count was tracked, allowing individuals to play with their overall activity. In addition to offering the possibility of direct intervention on people’s activity habits in natural context, the capacity of this mobile app to collect, in real-time, a large amount of objective real-world data can be useful for understanding the processes and outcomes of behavioral health interventions [23]. More specifically, these data can help make explicit when, where, for whom, and in what state for the participant, the intervention will produce the expected effect, notably thanks to continuous data collection over time. The within-person evolution in daily steps obtained via the app combined with between-person individual factors and intervention parameters is of great interest in this perspective.

Thus, the objectives of this study were to analyze the data collected in order (1) to examine within-individual evolutions of daily steps before, during, and after the intervention, (2) to test the effectiveness of a gamified program in real-life conditions on daily steps of participants versus nonparticipants, and (3) to explore the variables that could explain heterogeneity in response to the intervention. Based on previous results on gamification [20], we first hypothesized that daily steps would increase during and after the gamified program compared to baseline (H1). Second, we hypothesized that this improvement will be greater for participants than for non-participants (i.e., participants who registered on the app but did not complete any game, H2). Finally, we expected that the intervention’s characteristics (i.e., type and number of games), the context within the intervention was performed (i.e., population and settings), and the exposure to the intervention (i.e., engagement of participants with the app) will moderate the intervention effect (H3).

## Methods

### Study design and participants

This study retrospectively analyzed data from adult participants who had registered for a Kiplin program and had given consent for their data to be collected. To be included, participants must be 18 years old or older, have registered on the app between January 1^st^, 2019, and January 2^nd^, 2022, and logged daily steps (measured via their smartphone or an activity monitor) on a time frame of at least 90 days with less than 20% of missing daily observations. Of the 134,040 individuals who registered on the Kiplin app on this timespan, 4819 met the eligibility criteria. See Figure 1 for the study flow chart.

**Figure 1.**
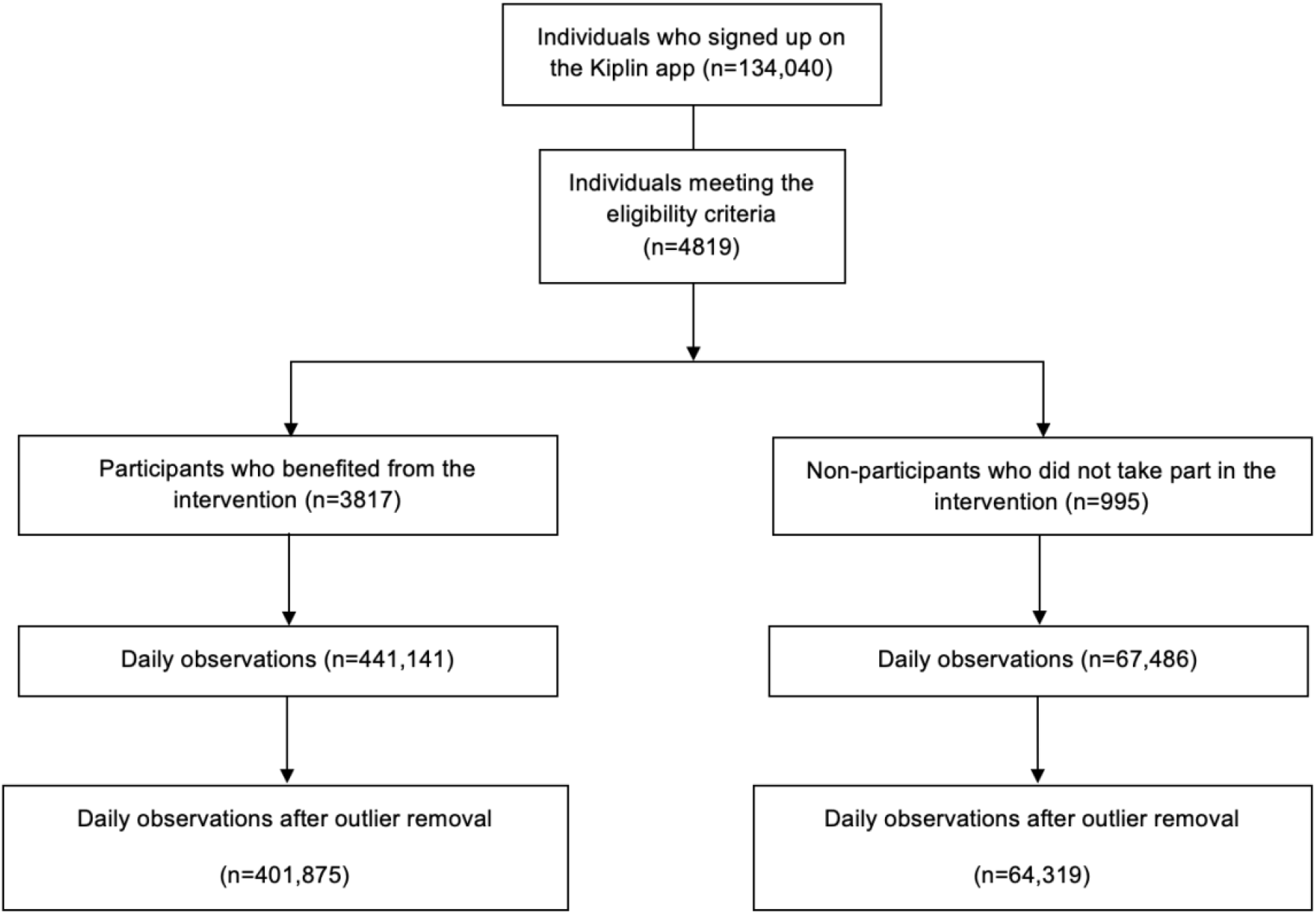
Study flow chart and screening of the databases.

Non-wear days were defined as days with fewer than 1000 steps and considered as missing observations – as previous research suggested that daily step values less than 1000 may not represent full data capture [24,25]. Days before the first day of the first game were considered as ‘baseline’ (14 days ± 42.9, median ± standard deviation), the period between the first day of the first game and the last day of the last game as ‘intervention period’ (19 days ± 31.2), and the days after the last day of the last game as ‘follow-up’ (90 days ± 22.8). We restricted the follow-up periods to 90 days post-intervention (i.e., 3 months).

Participants could receive the Kiplin intervention a) in the context of their work (i.e., primary prevention with employees), b) in a senior program (i.e., primary prevention with volunteer retirees), or c) as part of their chronic disease care (i.e., patients mainly treated for obesity or cancer). In all the aforementioned conditions, the program was paid not by the participant but by their employer or health care center.

Some participants registered for the program, created their account, but did not take part in the intervention (i.e., did not completed any game). These individuals were considered “nonparticipants” and were used as a control group (as proposed in previous research [26]). Similarly, the baseline period of these nonparticipants corresponds to the days prior to the date they were supposed to start the intervention period.

The study was approved by local Ethics Committee (IRB00013412, CHU de Clermont Ferrand IRB #1, IRB number 2022-CF063) with compliance to the French policy of individual data protection.

### The Kiplin intervention

The Kiplin intervention proposes time-efficient collective games accessible through an Android or iOS app. In all games, participants’ daily step counts are converted into points, allowing progression within the games. The Kiplin app retrieves participants’ daily step counts by integrating with the API (Application Programming Interface) of the applications used by participants to track their activity (such as Apple Health for iPhone users, Google Health for Android users, Garmin Health, etc.). In this way, participants could connect a wearable if they already owned one. Additionally, participants have access to a visual tool to monitor their daily and weekly step counts and to a chat for communication with other participants. Depending on the program, participants were offered one or several games lasting approximately 14 days each. If several games were proposed, these games followed each other in an interval of fewer than 60 days.

Participants could take part in four different games, with no option for selection. In “The Adventure,” the objective is to reach step goals collectively to progress towards a final destination. Players can track their progress on a map with checkpoints representing distances between different cities of a digital world tour (Figure 2A). In “The Mission,” participants engage in physical activity and collective challenges to unlock clues and attempt to solve missions (Figure 2B). In “The Board Game,” participants take on the role of forest rangers tasked with extinguishing fires. Achieving step goals allows progress on the board and advancement to higher levels, ultimately aiming to extinguish all fires and save forest residents (Figure 2C). Finally, in “The Challenge,” players aim to achieve the highest number of steps and complete challenges to earn trophies for their team. Team and individual rankings are available (Figure 2D).

**Figure 2.**
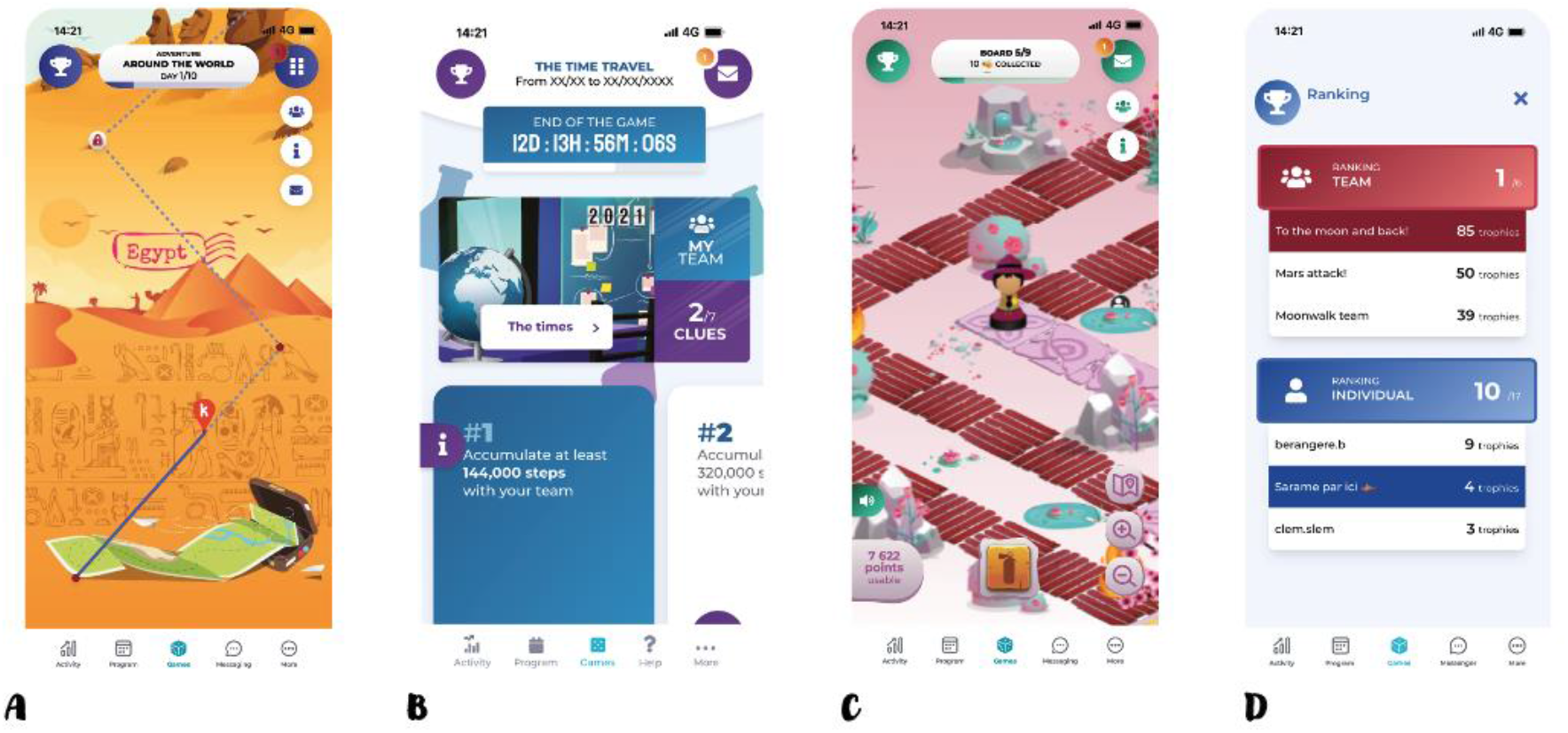
Screenshots of the Kiplin games. (A) The adventure. (B) The mission. (C) The Boardgame. (D) The Challenge.

These games include a multitude of gamification mechanisms such as points, trophies, leaderboards, a chat, challenges, narratives – mechanics that are closely linked to proven behavior change techniques [27]. Table 1 gives an overview of the gamification strategies included in the Kiplin games following the taxonomy proposed by Schmidt-Kraepelin et al. [28] and the associated behavior change techniques. While the games share common characteristics (e.g., collective gameplay, in-game challenges), it is important to note that the adventure and the challenge emphasize competition more than the others.

**Table 1.** Analysis of the game mechanics and content of the Kiplin app and associated behavior change techniques following Schmidt-Kraepelin et al. and Michie et al. taxonomies [27,28].

| Dimension | Characteristic | Gamification techniques | Associated Behavior change techniques |
| --- | --- | --- | --- |
| Gamification concept-to-user communication | Mediated | The gamification concept communicates with the user through an avatar named Pilot Kiplin (i.e., a real Kiplin team member animating the app who takes the persona of a funny mascot). Pilot Kiplin launches in-game challenges, announces results, and delivers internal messages aimed at motivating participants. These messages include tips to plan and implement PA in daily life and information on the benefits of walking on health. | Instruction on how to perform a behavior (4.1)<br>Information about health consequences (5.1) |
| User identity | Static self-selected identity | Participants are only able to select a nickname and personalize the name of their team. |  |
| Rewards | Internal | Rewards that participants can earn through the app are solely internal and virtual, such as points and trophies. | Feedback on behavior (2.2)<br>Self-monitoring of behavior (2.3)<br>Cue signaling reward (7.2)<br>Behavioral practice/rehearsal (8.1)<br>Non-specific reward (10.3) |
| Competition | Direct and indirect | Participants can compete directly via in-game challenges (i.e., at specific intervals, a competitive challenge commences between teams. To secure victory and earn a trophy, one team must accumulate more points or steps than their rival team within the allotted timeframe) or indirectly via the point system and leaderboards. These challenges are announced in advance, encouraging players to plan their activities so as to be active on the day of the battle. | Action planning (1.4)<br>Feedback on behavior (2.2)<br>Social comparison (6.2) |
| Target group | Patients and healthy individuals | The app can be offered to patients in clinical settings, as well as in preventive initiatives among employees or seniors. |  |
| Collaboration | Cooperative | All games operate on a collaborative basis: participants are organized into teams and must collectively complete challenges to advance or win. They can use the chat feature to communicate, support each other, and exchange ideas with their teammates. | Social support (unspecified, 3.1)<br>Social support (emotional, 3.3) |
| Goal-setting | Externally set | Goals, cutoffs, and the number of steps required to earn a trophy are predetermined by the app developer or the healthcare professional. | Review behavior goals (1.5)<br>Discrepancy between current behavior and goal (1.6) |
| Narrative | Episodal | Narratives are unlocked as participants reach new milestones, such as unlocking a new clue in the mission, reaching a higher board level in the board game, or reaching a new city in the adventure. |  |
| Reinforcement | Positive | The app focuses solely on highlighting current and future successes, aiming to avoid stigmatizing users based on their initial inactivity or health status. |  |
| Persuasive intent | Behavior change | The app aims to enhance individuals' daily step counts through game mechanics. |  |
| Level of integration | Inherent | To participate in the games, individuals must engage in physical activity. Progression within the game is contingent upon the steps performed; without activity, players cannot advance. |  |
| User advancement | Presentation only | The progression is presented through the clues collected in the mission, to the progression to different cities in the adventure and to new board levels in the boardgame. This structure encourages participants to engage in and repeat the target behavior. | Graded tasks (8.7)<br>Reward approximation (14.4) |
| Other tools of the app |  | Self-monitoring tool, notifications | Self-monitoring (2.2)<br>Prompt/cues (7.1) |

### Variables

The variables of interest were selected on the basis of the behavior change intervention ontology of Michie et al. [22] and included (1) the longitudinal evolution of daily steps, (2) the exposure of each participant to the intervention, (3) the intervention parameters, and (4) the context (participants’ characteristics and settings), as these variables are likely to influence the intervention effect. Table 2 specifies the measures of interest and their operationalization.

**Table 2.**
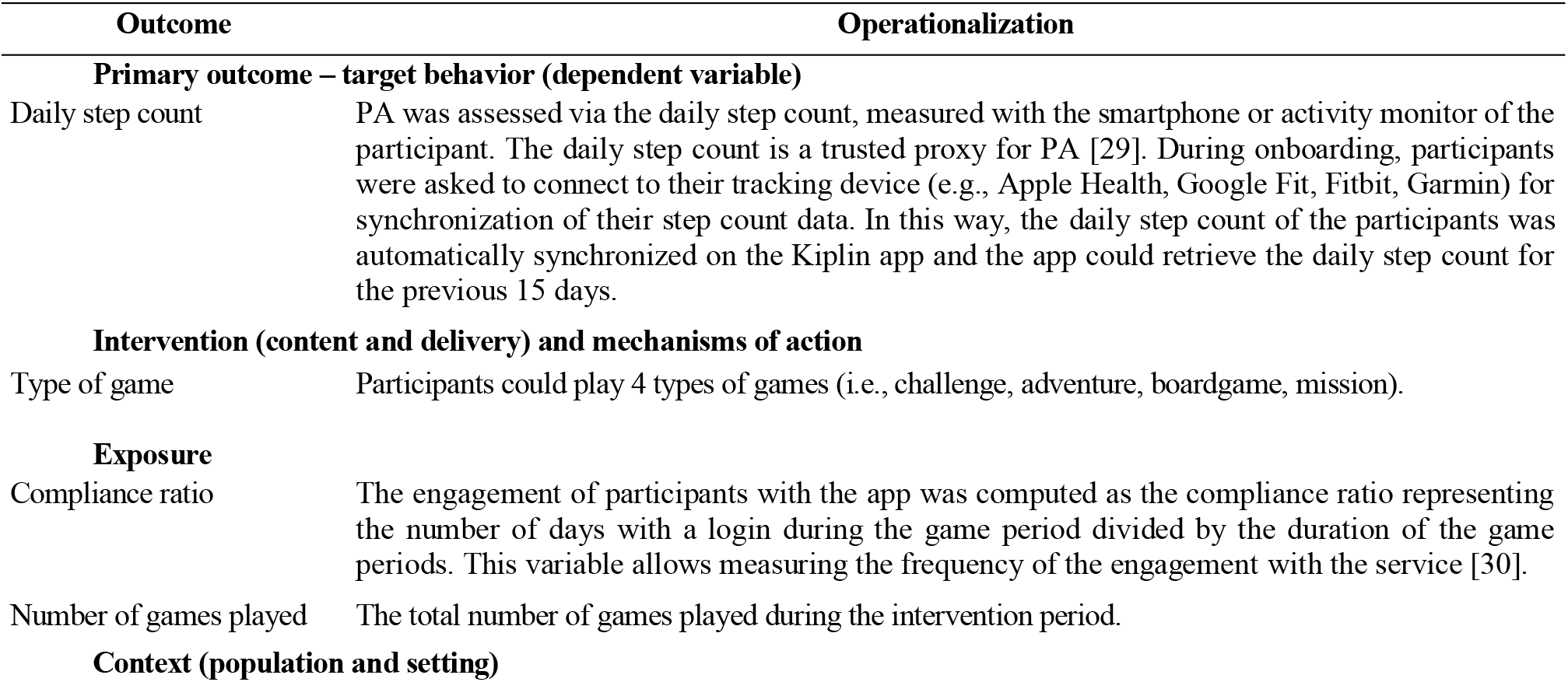
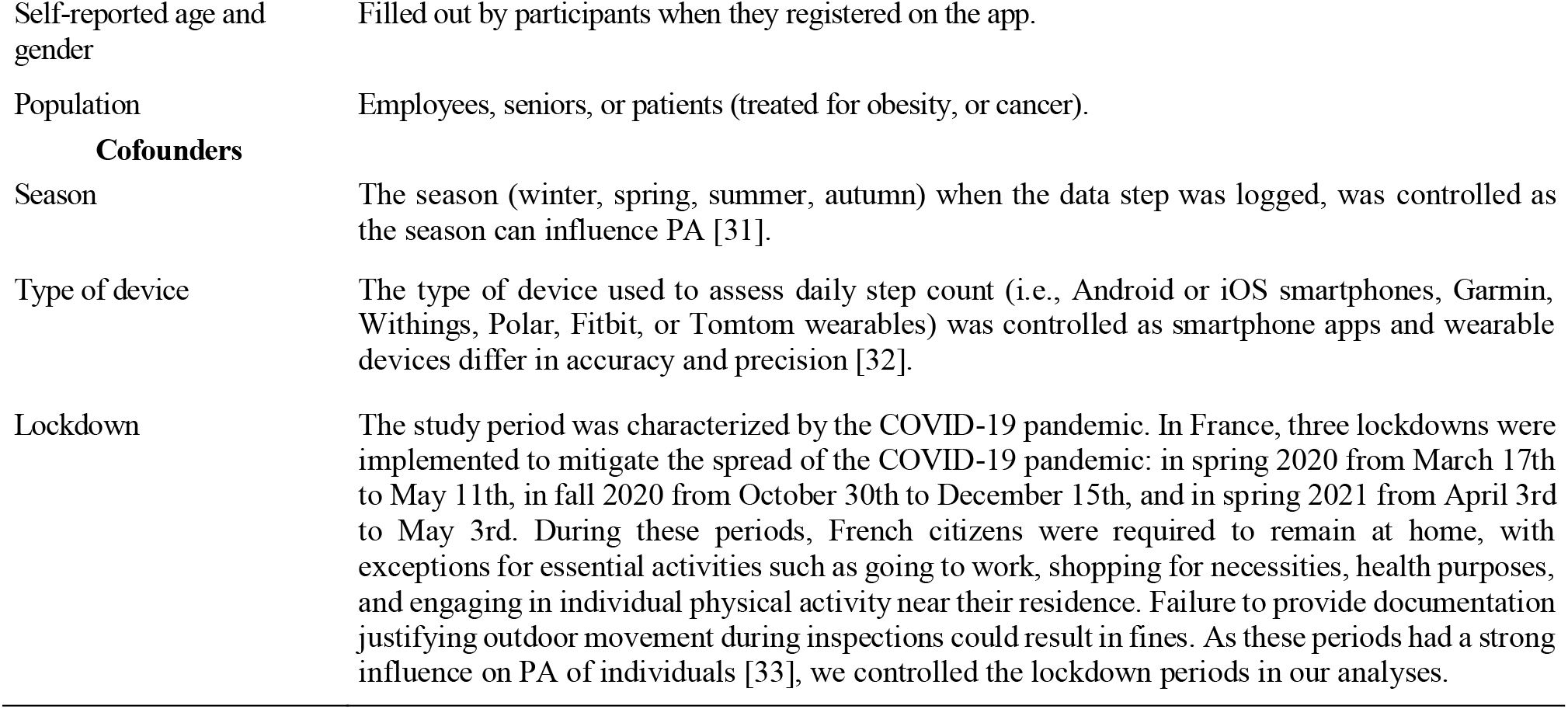
Operationalization of the variables.

### Statistical analyses

We calculated the step count increase by subtracting the baseline average daily step count from the average daily step count during the intervention or follow-up periods for each participant and then computed the relative change in %.

Mixed-effects models were used to 1) analyze within-person evolution across time (i.e., changes in daily steps between baseline, intervention, and follow-up periods), and across participants and nonparticipants, and 2) examine the associations between intervention parameters, exposure to the intervention, participants’ characteristics and settings, and the daily steps evolution. This statistical approach controls for the nested structure of the data (i.e., multiple observations nested within participants), does not require an equal number of observations from all participants [34], and separates between-person from within-person variance, providing unbiased estimates of the parameters [35,36].

First, an unconditional model (i.e., with no predictor) was estimated for each variable to calculate intra-class correlations (ICC) and estimate the amount of variance at the between and within-individual levels, which allowed us to determine whether conducting multilevel models was relevant or not. Then, a model that allowed random slope over time (i.e., model with random intercept and random slope) was compared to the null model (i.e., with only random intercept) using an ANOVA, to evaluate whether the less parsimonious model explain a significantly higher portion of the variance of the outcome, compared to the unconditional model [37,38]. Third, between-level predictors and confounding variables were added to another model (Model 1)*^1^ and compared to the previous models. Finally, intervention characteristics as well as their interactions with the phases (i.e., baseline/intervention/follow-up) of the study were added in a final model excluding nonparticipants (Model 2)^*2^.

Model fit was assessed via the Bayesian Information Criterion (BIC) and −2-log-likehood (−2LL) [39]. All models were performed using the *lmerTest* package in the R software [40]. An estimate of the effect size was reported using the marginal and conditional pseudo R^2^. When the interaction terms turned significant, contrasts analyses were computed using the *emmeans* package [41]. Models’ reliability (estimated with residual analyses) and outliers detection were performed using the *Performance* package [42]. In addition to subtracting non-wear days (defined above), we removed outliers via the ‘check_outliers’ function [42] that checks for influential observations via several distance and clustering methods (i.e., Z-scores, Interquartile range (IQR); Equal-Tailed Interval). Sensitivity analyses were conducted using all data (including data before outlier imputation) and are available in supplementary materials.

The data and code for the statistical analyses used in the present study are available on Open Science Framework [43].

## Results

### Descriptive Results

Descriptive results are presented in Table 3. The final sample included 4819 adults (mean age = 42.7 ± 11.5 years; 60% women). Participants wore an activity monitor measuring their daily step count for an average of 113 days (range = 90 – 686 days). A total of 34,922 daily steps observations were missing (i.e., daily data missing or considered as a non-wearing day), which is equivalent to 6.4% of missing data on the full dataset.

**Table 3.** Descriptive statistics.

|  | Participants | Nonparticipants |
| --- | --- | --- |
|  | N=3817 | N=995 |
| *Sociodemographics* |  |  |
| Age, mean (SD) | 43.2 (11.08) | 41.0 (12.81) |
| Female (%) | 2313 (62) | 510 (53) |
| Employees (%) | 3,526 (92) | 978 (98) |
| Patients (%) | 194 (5) | 17 (2) |
| Seniors (%) | 97 (2) | - |
| *Exposure* |  |  |
| Compliance ratio, mean (SD) | 0.84 (0.23) | 0 |
| Games played, mean (SD) | 1.28 (0.9) | 0 |
| In-game days, mean (SD) | 22.06 (16.24) | 0 |
| *Observations in each type of game* |  |  |
| Adventure (% of all days) | 21,316 (33) | - |
| Boardgame (% of all days) | 4,093 (6) | - |
| Challenge (% of all days) | 32,801 (50) | - |
| Mission (% of all days) | 6,915 (11) | - |
| *Type of device used* |  |  |
| Android smartphone (%) | 1076 (28) | 286 (29) |
| iOS smartphone (%) | 810 (21) | 533 (54) |
| Fitbit (%) | 750 (20) | 52 (5) |
| Garmin (%) | 1,071 (28) | 109 (11) |
| Polar (%) | 5 (0.1) | - |
| Tomtom (%) | 3 (0.08) | - |
| Withings (%) | 90 (2) | 9 (1) |
| *Observations in each season* |  |  |
| Winter (% of all days) | 110,517 (24) | 17,451 (24) |
| Spring (% of all days) | 94,961 (20) | 21,162 (30) |
| Summer (% of all days) | 129,039 (27) | 8,804 (12) |
| Fall (% of all days) | 138,429 (29) | 24,086 (34) |
| *Observations in each lockdown* |  |  |
| 1<sup>st</sup> lockdown – spring 2020 (% of all days) | 10,872 (3) | 925 (1) |
| 2<sup>nd</sup> lockdown – fall 2020 (% of all days) | 32,298 (8) | 4,110 (6) |
| 3<sup>rd</sup> lockdown – spring 2021 (% of all days) | 23,435 (6) | 1,757 (2) |

We tested for statistical differences in sociodemographic variables and baseline daily steps between participants and nonparticipants using t-tests and Chi^2^ tests. Results revealed significant differences for age (*t* = −6.9149, *p*<.0001), gender (χ^2^ = 4028.3, *p*<.0001), and baseline daily steps (*t* = −19.75, *p*<.0001). However, in large samples, *p*-values may drop below the alpha level despite effect sizes that are not practically meaningful [44]. Therefore, we mainly examined the magnitude of the effect sizes of these differences and observed very small to small effects (*d* = −0.03 for age, *d* = −0.17 for baseline daily steps, and *w* = 0.09 for gender). According to Magnusson [45], the interpretation of these effect sizes suggests that, for age and baseline daily steps, approximately 98.8% and 93.2% of individuals in both groups overlap, respectively. Additionally, there is approximately a 50.8% and 54.8% chance that a randomly selected individual from the nonparticipant group would have a higher score than a randomly selected individual from the participant group. Therefore, we considered that the differences are minor between the two groups. Finally, these variables were controlled in our mixed-effects models as they were included as fixed effects.

### Is the gamified program effective to promote PA? (H1)

During the intervention period, participants increased their daily steps by 2619 steps per day on average (+55.6%), compared to the baseline period, and by 317 steps per day on average during the follow-up period (+13.8%), compared to the baseline. In comparison, the daily step count of the control group remained more or less stable throughout the same timeframe with a mean increase of 151 daily steps compared to baseline (+7.5%).

Overall, contrast analyses of the model for the intervention participants (Model 2, Table 4) revealed a negative effect of the intervention on the daily step count during the intervention phase compared to baseline activity (*b* = −0.09, 95 CI [-0.14; −0.05], *p*<.0001) and no significant effect (*b* = 0.01, 95 CI [- 0.05; 0.06], *p*=0.79) during follow-up periods compared to baseline. However, the patterns were different when participants were stratified by baseline PA. Participants with lower baseline daily steps (<5000 steps per day or 5001-7500 steps per day) showed a significant increase of their daily steps during the intervention and the follow-up, both compared to the baseline (respectively *b* = 0.25, 95 CI [0.22; 0.28], *P*<.0001 and *b* = 0.12, 95 CI [0.09; 0.15], *p*<.0001).

**Table 4.**
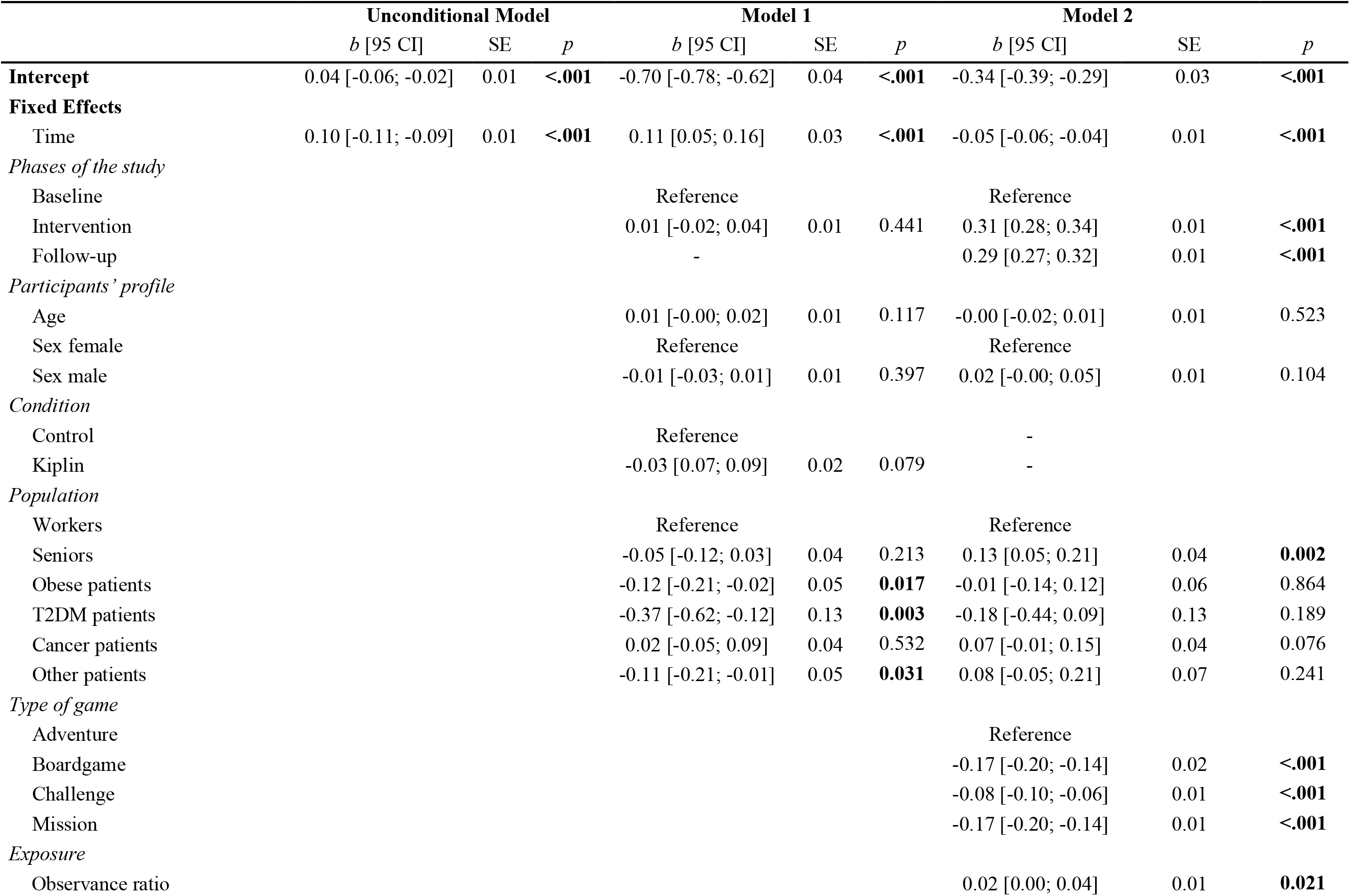
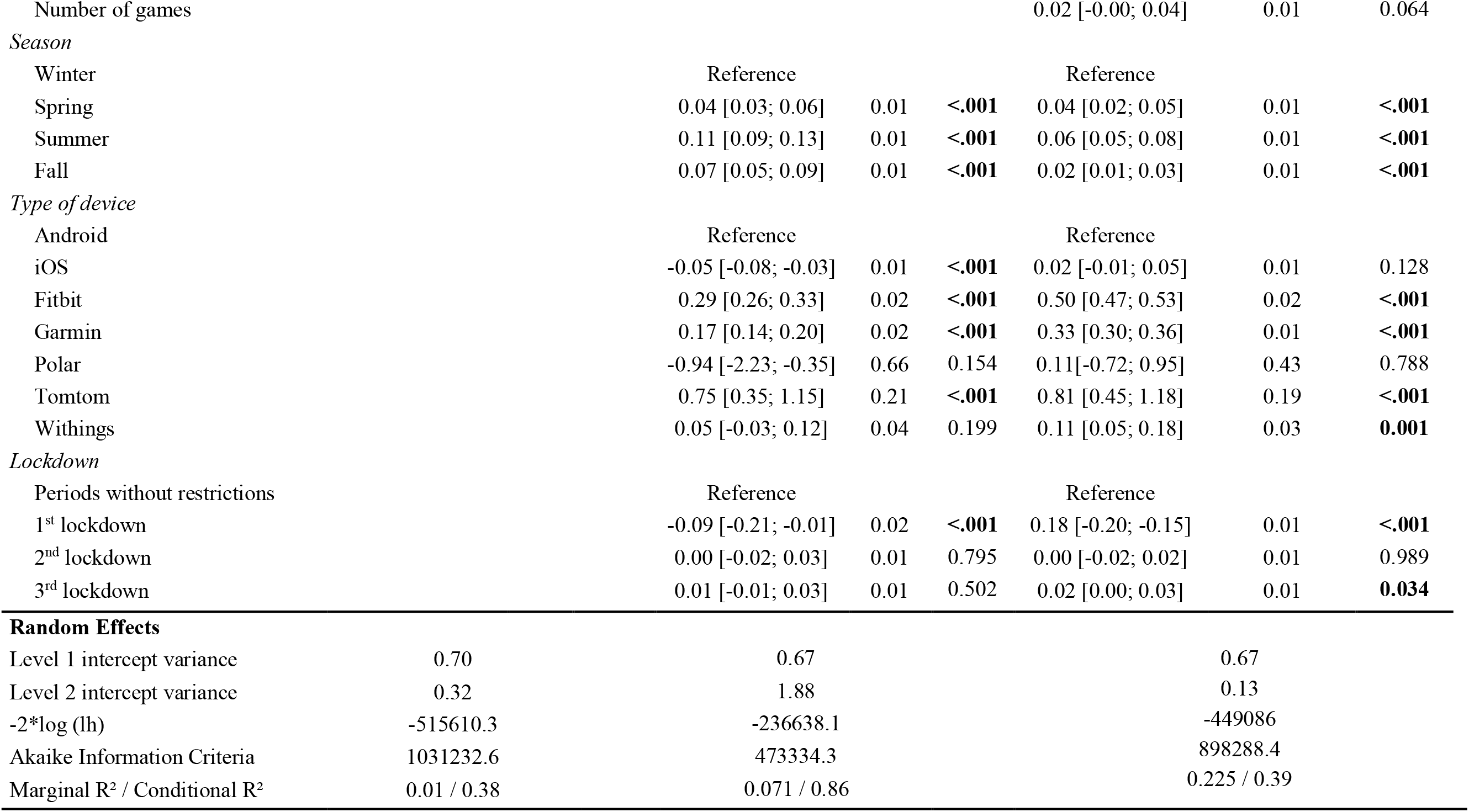
Mixed effect models’ estimates.

Participants with initial values between 7501 and 10000 steps did not have a significant increase in their daily steps during the intervention (*b* = 0.00, 95 CI [- 0.05; 0.05], *p*=0.99) nor during the follow-up period (*b* = −0.01, 95 CI [-0.04; 0.02], *p*=0.44), compared to baseline. Participants who performed more than 10000 baseline steps had significant deteriorations during the intervention (*b* = −0.13, 95 CI [-0.19; −0.08], *p*<.0001) and follow-up (*b* = −0.06, 95 CI [-0.10; −0.03], *p*=0.0001). These trends are depicted in Figure 3 and in Table 5. Results were similar in sensitivity analyses that used data without outlier imputation, except for participants with initial daily step count between 7501 and 10000 daily steps who observed significant improvements during and after the intervention (Table S1 and S2 in Multimedia appendix 1).

**Table 5.**
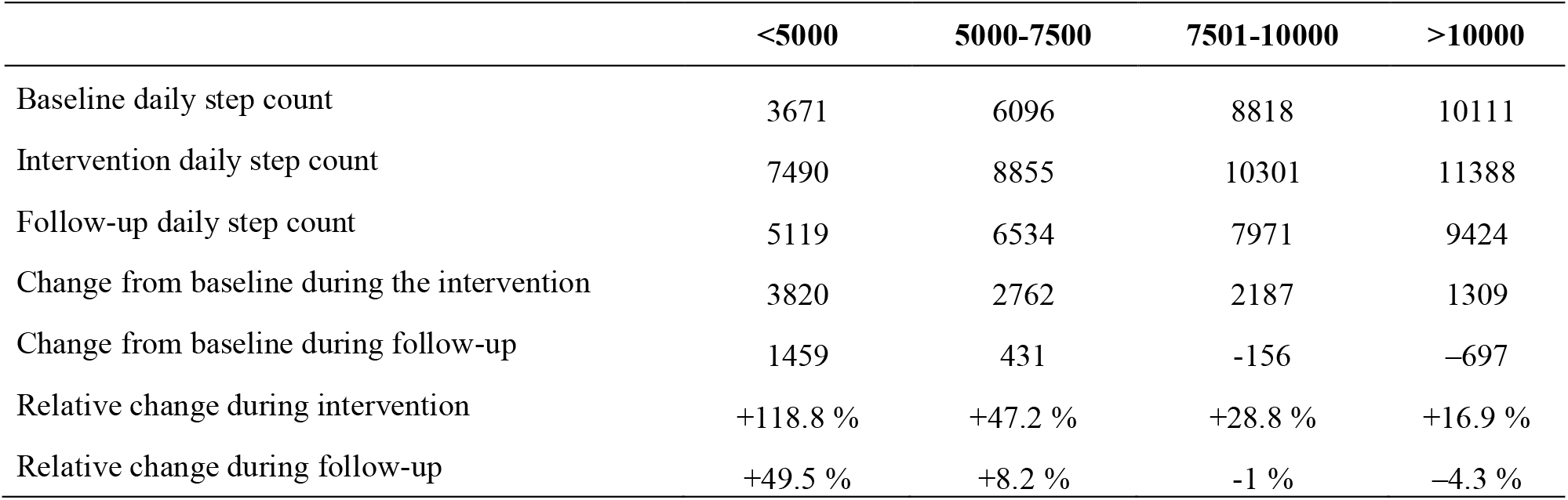
Description of the mean daily step count during baseline, intervention, and follow-up periods, changes, and relative changes from baseline in function of participants’ baseline daily step count.

**Figure 3.**
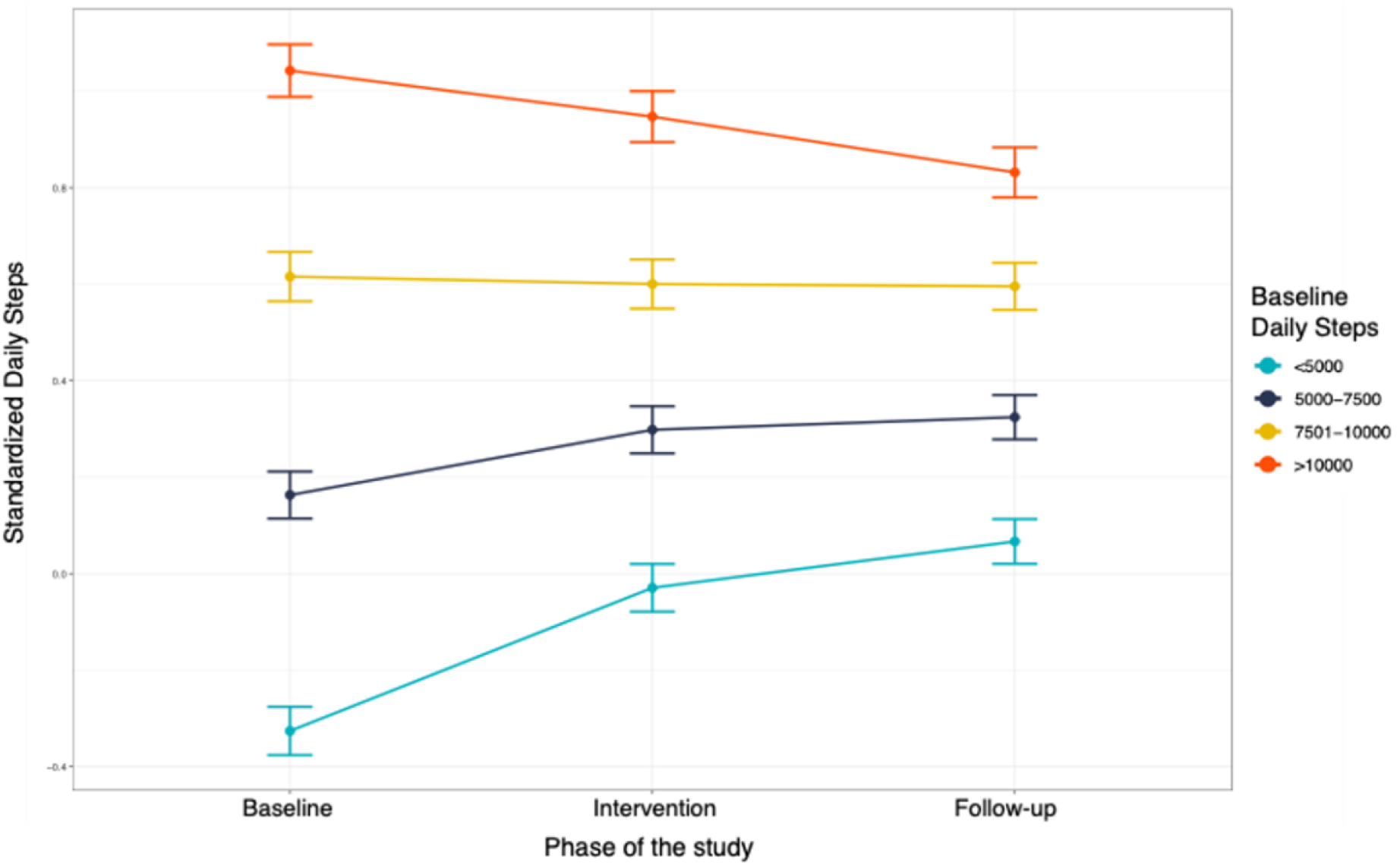
Changes in daily steps throughout the study phases for participants who received a Kiplin program, stratified by baseline daily steps

**Figure 4.**
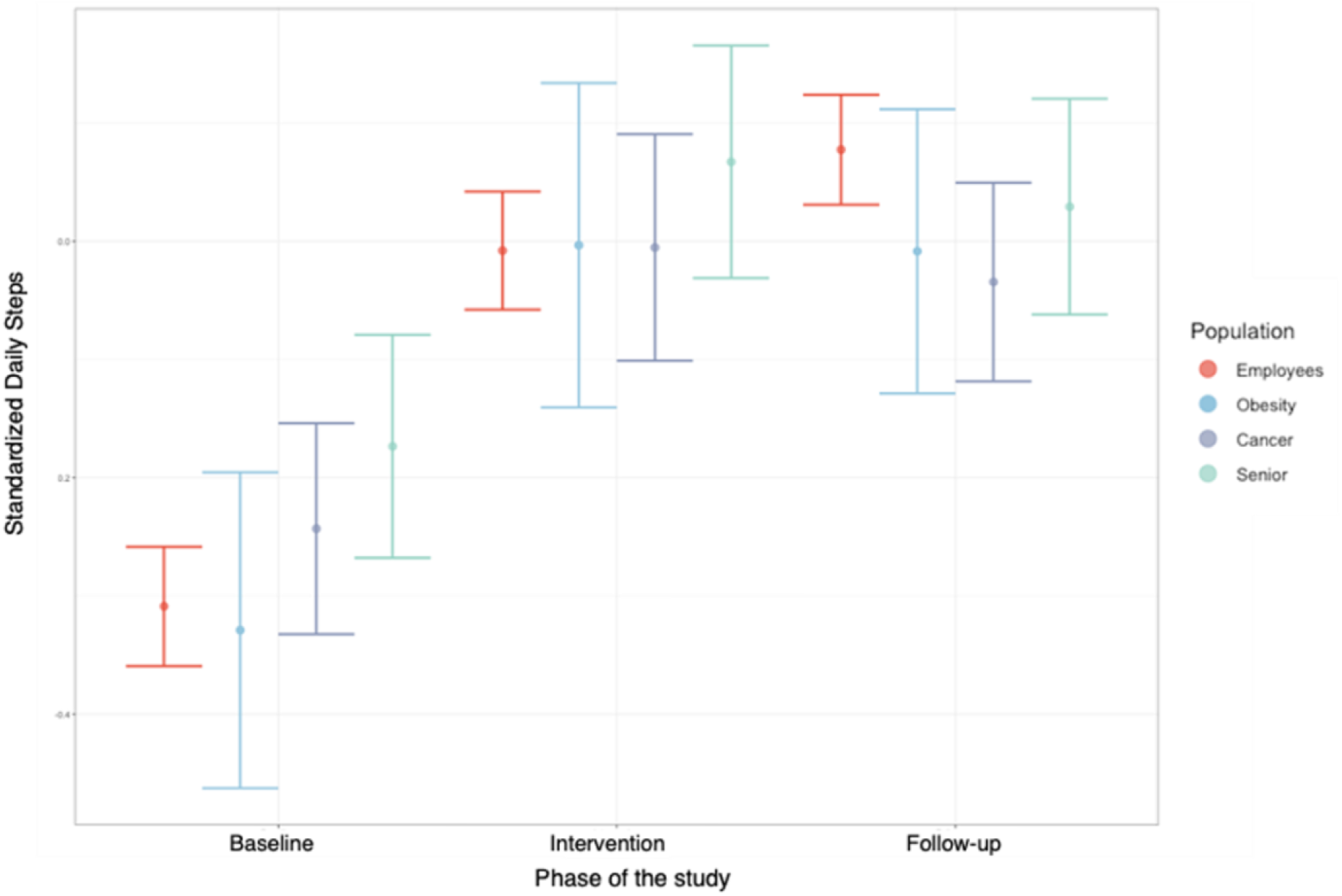
Changes in daily steps across the study phases for the different populations who received a Kiplin program.

In parallel, contrast analyses comparing the effectiveness of the Kiplin intervention who used smartphones to collect their daily steps in comparison to participants who used a wearable showed a significantly greater effect among smartphone users, both during the intervention phase (*b* = 0.09, 95 CI [0.07; 0.11], *p*<.0001) and the follow-up period (*b* = 0.04, 95 CI [0.01; 0.06], *p*=0.001). These results are illustrated in Figure S1 in Multimedia Appendix 1.

### Is the intervention effect greater for participants compared to nonparticipants? (H2)

In Model 1 (Table 4), participants who received the Kiplin intervention had a significantly greater increase in mean daily steps between baseline and the intervention period, compared with nonparticipants (*b* = 0.54, 95%CI [0.52; 0.58], *p*<.0001). The results were similar in sensitivity analyses (Table S2 in Multimedia appendix 1).

### What are the moderators of the intervention effect? (H3)

The Model 2 estimates are displayed in Table 4. The variables under consideration explained 39% of the variance in daily steps. In this model, we tested the hypothesized interactions, to investigate predictors associated with the efficiency of the intervention (Table S4 in Multimedia appendix 1). Contrast analyses were conducted on significant interactions and revealed that the age (*b* = 0.05, *p*<.0001) and the compliance ratio (*b*=0.37, *p*<.0001) were positively associated with the change in daily steps between baseline and intervention. Specifically, the older the age, the more regularly the individuals played and the more effective the intervention was. On the other hand, the number of games played by participants was negatively associated with this change (*b* = −0.02, *p*=0.02). In other words, the longer the intervention and the higher the number of games, the less effective the intervention. For categorical outcomes, contrast analyses revealed differences in the intervention effect between the different populations. Compared to employees, patients treated for cancer (*b* = −0.18, *p*<.0001), and seniors (*b* = −0.19, *p*<.0001) observed a significantly weaker effect of the intervention in comparison to baseline PA. There was no significant difference between employees and patients treated for obesity (*b* = −0.07, *p*=0.13). All the results of these analyses are available in supplementary materials (Multimedia appendix 1).

Finally, Model 2 estimates revealed that participants were significantly more active in the adventure and the challenge compared to the boardgame and the mission (Table 4).

## Discussion

This study demonstrated a significant increase of daily steps among participants engaging with Kiplin intervention compared to nonparticipants over the same period. Interestingly, the intervention effect varied in function of the baseline daily step count of individuals. Participants with lower baseline steps (<7500 steps per day) significantly improved their PA both during the intervention (between +34% and +76%) and follow-up periods (between +10% and +33%) whereas participants with more than 7500 steps had no significant change or significant decreases.

These results suggest that a gamified program is more efficient for inactive individuals compared to active ones, with the existence of a plateau effect. They also support recent findings [20,46] and the ability of gamified interventions to improve daily steps both during and after the end of the program and in real-life settings [47] – at least for the more inactive individuals. This efficacy is noteworthy given the challenges faced by current behavioral interventions in promoting PA in the long haul [9].

The SDT offers a valuable framework for elucidating the disparate outcomes observed among initially active and inactive participants. Whereas gamification strategies could enhance the autonomous motivation of inactive participants as suggested by a previous study [48], the use of rewards on already motivated people could undermine this motivation. Known as the overjustification effect [18], this phenomenon suggests that if people receive rewards for doing an activity that they used to enjoy, they are likely to discount the internal reason, and thus become less intrinsically motivated than before getting the rewards. This could explain why the same intervention had positive effects on inactive participants who performed more daily steps after the end of the intervention (i.e., during follow-up periods) compared to already active ones who observed significant decreases after the intervention compared to their baseline daily steps.

Moreover, results indicating that the intervention is more effective among users who use their smartphones to track their step counts with the Kiplin app, compared to those who already own and use a wearable device – and are significantly more active at baseline – further reinforce this argument. Individuals who already possess an activity monitor are likely motivated to monitor their daily steps, potentially diminishing the additional impact of gamification rewards. Consequently, the introduction of gamification may have less influence or even produce counterproductive effects on their behavior, particularly when compared to those who solely rely on their smartphones for activity tracking in the context of the intervention.

The results of this study also stressed that older age may not be incompatible with gamified interventions. Indeed, the intervention effectiveness was moderated by the age of the individual and gamification was more efficient among older individuals, compared to younger ones. These findings are in line with a previous study [49] which reported higher utilization of gamification features among older users. The authors postulated that older adults pay generally more attention to their health and thus have a stronger intention to engage in a health program. From another perspective and in light of the gamification strategies embedded in the Kiplin intervention, these results could also be explained by the fact that these strategies are accessible – inspired by traditional board game rules and mechanics widely known in the general population – and thus may be more attractive for older populations. Prior research suggested that the most engaging game mechanics may diverge between youth and other populations [50], and we can expect that younger populations may prefer more complex game mechanics and need more novelty during the intervention to stay interested by the service.

Regarding the effects of the gamified intervention according to the characteristics of the population, a stronger effect was found for programs among employees and patients treated for obesity. While these results warrant caution due to the variability observed in patients or senior participants, these findings suggest that gamified interventions are suitable for both primary and tertiary prevention, as suggested by previous work [20].

The findings of this study also offer valuable insights that could help to improve future intervention design. Firstly, exposure to the content is essential for the gamified intervention to be effective. It is interesting as gamification has often been assimilated into a self-fulfilling process permitting automatic engagement of participants. These results are consistent with previous findings demonstrating that higher use of gamification features was associated with greater intervention effectiveness [49,51]. If gamification can ultimately increase program engagement, developers need first to design their apps to be as attractive as possible and optimize retention.

Secondly, the results revealed that the total number of games played was negatively associated with the intervention effect, suggesting that shorter interventions could be more beneficial for behavior change. These results are in line with previous research [20,52] suggesting that digital interventions shorter than 3 months tend to yield greater benefits. It also suggests a « dose-response » relationship in inverted U shape, with an optimal “middle” to find. Nevertheless, it is important to consider that Kiplin programs incorporating multiple games are built in such a way as to decimate several doses at regular intervals. Periods without games were therefore considered in the intervention phases and could explain why, overall, the shorter games were more efficient. More refined analyses of the intervention effect over time will be necessary in the future.

Thirdly, the daily step count of participants was significantly higher in the adventure and the challenge. These two games are characterized by their competitive nature, placing a stronger emphasis on leaderboards compared to the other two games, which are more centered on collaboration. In this vein, Patel et al. [53] observed that the competitive version of their gamified intervention outperformed the collaborative and supportive arms. Moreover, various studies have highlighted that leaderboards are a particularly successful gamification mechanic [49,54].

### Strengths and limitations

This study has several strengths, including its large sample size, the intensive objective PA measurement in real-life conditions through daily steps, and the longer baseline and follow-up duration compared with most trials on gamification that typically incorporate measurement bursts dispersed across time [20]. However, several limitations should be considered. First, this study was observational and not a randomized controlled trial. Thus, we cannot establish the causality of the intervention’s effect on outcome improvement. The nonparticipants are not a true control group. If they did not receive the intervention, it may be because they were unable to join or for underlying motivational reasons that could impact their PA. Second, intervention lengths differed between participants. Third, if mixed-effects models are useful for describing trends in PA behavior change over time, they are limited in their capacity to assess precise fluctuations patterns of non-stationarity behavior such as daily step counts [55] across time. Future longitudinal studies could benefit from employing time series analyses to more accurately describe these patterns of change. Finally, the compliance ratio used in this study as a proxy for engagement tends to oversimplify the exposure of participants to the service. Complementary measures of behavioral engagement (e.g., using the number of logins, time spent per login, and the number of components accessed) and affective engagement (e.g., emotions, pleasure) should be considered to draw the longitudinal impact of the engagement of the participants on the intervention effect.

## Conclusion

In this study, we conducted a comprehensive analysis of real-world data from over 4800 individuals, suggesting the impact of a gamified intervention in real-life settings. Our findings indicate that the Kiplin intervention led to a significantly greater increase in mean daily steps from baseline among user compared to nonparticipants. Interestingly, responses to the intervention were significantly different as a function of individuals’ initial daily step counts. Whereas participants with less than 7500 baseline daily steps had significant improvements both during the intervention and follow-up periods with +3291 daily steps during the program and +945 after the intervention on average, the intervention had no effect on participants with initial values >7500. The motivational effect of gamification could therefore depend on the initial PA and motivational profile of the participants. This result can also be interpreted in light of our observation that participants who already owned a wearable, and thus were likely already motivated to engage in PA, exhibited significantly lower effects compared to less experienced participants who utilized their smartphones to track their step counts. This study also revealed that the age of participants and their engagement with the app were positively and significantly associated with the intervention effect while the number of games played was negatively associated with it.

Overall, the results of this study suggest that gamification holds promise in promoting the daily steps of inactive populations, with demonstrated short and medium-term effects. Importantly, this study represents a pioneering effort as one of the first to examine the longitudinal effect of a gamified program outside the context of a trial, using intensive real-world data. As such, the findings are quite generalizable in similar settings and reaffirm the value of gamification in both primary and tertiary prevention efforts across a diverse range of age groups.

## Data Availability

The data and code for the statistical analyses used in the present study are available on Open Science Framework (https://osf.io/scnu7/).

## Additional Information

### Contributions

Conceptualization of the study: AM; data curation: AM, GH; investigation: AM; methodology and statistics: AM, CF; software: AM; writing – original draft: AM; writing – review & editing: all the authors.

### Data & Code Accessibility

The anonymized data used in this study and the R code are available on the Open Science Framework (https://osf.io/scnu7/).

## Footnotes

* 1 The equation for the Model was the following: Y_ij_ = (β_0_ + γ_0i_ + θ_0j_) + (β_1_ + θ_1j_) Time_j_ + β_2_ Phase_j_ + β_3_ Age_j_ + β_4_ Sex_j_ + β_5_ Population_j_ + β_6_ Season_j_ + β_7_ Captor_j_ + β_8_ Baseline PA_j_ + β_9_ Lockdown_j_ + β_10_ Condition_j_ × Phase_j_ + ε_ij_ where β_0_ to β_10_ are the fixed effect coefficients, θ0j and θ1j are the random effect for the participant j (one random intercept and one random slope), γ_0i_ is the random effect for the Time i (random intercept), and ε_ij_ is the error term.

*2 The equation for the Model was the following: Y_ij_ = (β_0_ + γ_0i_ + θ_0j_) + (β_1_ + θ_1j_) Time_j_ + β_2_ Phase_j_ + β_3_ Age_j_ × Phase_j_ + β_4_ Sex_j_ + β_5_ Population_j_ × Phase_j_ + β_6_ Season_j_ + β_7_ Captor_j_ + β_8_ Baseline PA_j_ × Phase_j_ + β_9_ Lockdown_j_ + β_10_ Compliance ratioj × Phasej + β11 Number of games playedj × Phasej + β12 Type of Game_j_ + ε_ij_ where β_0_ to β_12_ are the fixed effect coefficients, θ_0j_ and θ_1j_ are the random effect for the participant j (one random intercept and one random slope), γ_0i_ is the random effect for the Time i (random intercept), and ε_ij_ is the error term.

